# Risk of Inflammatory Bowel Disease in Psoriasis Patients Treated with Anti-Interleukin-17 Agents: A Bayesian Metaanalysis

**DOI:** 10.1101/19012179

**Authors:** N. Gill, M. Pietrosanu, R. Gniadecki

## Abstract

**Background:** Use of interleukin-17 inhibitors (IL-17i) in psoriasis has been associated with an increased risk of inflammatory bowel disease (IBD). However, the clinical significance of this association is not understood.

**Objectives:** To quantify the absolute risk of IBD in patients with psoriasis treated with IL-17i, stratified by known IBD risk factors.

**Methods:** Literature searches were performed to identify known IBD risk factors and the prevalences were quantified by a meta-analysis of proportions. The Bayesian model was used to estimate the probability of a new-onset or a flare of IBD in patients with psoriasis.

**Results:** The prevalence of Crohn’s disease (CD) or ulcerative colitis (UC) in the general psoriasis population was 0.0010. Use of IL-17i increased the risk of CD to 0.0037 and UC to 0.0028, translating to a number needed to harm (NNH) of 373 for CD and 564 for UC. In patients who had concomitant hidradenitis suppurativa, the use of IL-17i was associated with a decrease in NNH for CD and UC to 18 and 76, respectively, whereas for patients with a family history of IBD, the NNH values were 6 (for CD) and 10 (for UC).

**Conclusions:** In patients with no risk factors, the probability of IBD flare or onset during IL-17i treatment is negligible and additional IBD screening procedures are not indicated. In contrast, the patients with psoriasis who have hidradenitis suppurativa or first-degree family history of IBD as risk factors should be monitored for signs and symptoms of CD and UC during IL-17i therapy.

## Introduction

Psoriasis is a chronic immune-mediated inflammatory skin disorder affecting 2-3% of the Western population.^1^ Interleukin-17A is a proinflammatory cytokine involved in the pathogenesis of psoriasis and is an effective therapeutic target.^2,3^ Secukinumab and ixekizumab are among the IL-17A inhibitors (IL-17i) currently approved in psoriasis and have shown superiority to placebo^4^ and other biologics^5^ with minimal adverse events. However, recent observations that IL-17i may exacerbate or even induce inflammatory bowel disease (IBD)^6^ led the Federal Drug Administration (FDA) to encourage caution when prescribing to patients with a history or risk of IBD.

IBD comprises Crohn’s disease (CD), characterized by transmural inflammation in the gut, and ulcerative colitis (UC) in which inflammation is limited to the mucosal layer of the colon. Common clinical symptoms and signs include abdominal pain and diarrhoea, sometimes with hematochezia. The prevalence of CD and UC in North America ranges between 96.3 - 318.5 and 139.8 - 286.3 per 100,000 persons, respectively.^7^ Distinguishing CD from UC is important due to differences in management and prognosis. Surgical resection of the colon and rectum is considered curative in UC^8^ whereas CD has a 10-year postoperative recurrence rate of 44– 55%.^9,10^ Ileocolonoscopy with ileal biopsies and imaging constitute the gold standard for diagnosis of IBD^9,11^ but an unequivocal classification may not be possible in up to 15% of cases.^10,12^ Endoscopy is not an efficient diagnostic technique in asymptomatic patients^11^ and there are no robust, non-invasive screening methods to assess the risk of subclinical IBD in the population. Use of faecal calprotectin is a sensitive test measuring disease activity but should not be used for screening purposes as it would lead to a vast overestimation of the risk of IBD.^13^

Thus, the dermatologist prescribing IL-17i is faced with the practical challenge of carrying out a simple yet cost-effective assessment of the risk of IBD in psoriasis patients. The current FDA warning for secukinumab^14^ and ixekizumab^14^ does not provide any guidance on appropriate screening procedures in patients who do not have a diagnosis of IBD. There is, therefore, a need for a quantitative and clinically understandable estimate of the risk of IBD in a patient treated with IL-17i. To meet this clinical need we used the Bayesian approach to provide risk estimates of IBD in psoriasis patients treated with IL-17i.

## Materials and Methods

### Search Strategy

We searched the Ovid MEDLINE database for papers reporting risk factors for IBD. The inclusion criteria were: adult patients (age > 18 years), report of an IBD risk factor with a statistically significant association, and a reported risk factor that can objectively be assessed for in the clinic. For example, studies assessing components of diet as risk factors were not considered as subjective and not readily reproducible. Subsequently, we performed separate searches for each individual risk factor to obtain the data on the prevalence of those risk factors. For the data in IL-17i treated patients, we pooled secukinumab and ixekizumab studies assuming that these drugs do not substantially differ in their ability to modulate IBD. Studies were considered for inclusion if they involved adult patients (age > 18), contained data required to calculate the prevalence of the given risk factor, reported original data and were written in English. Abstracts, unpublished data, and letters to the editor were also evaluated. Articles meeting the inclusion criteria after a title and abstract screen underwent full article review. The reference lists of all selected papers were further reviewed to identify additional potentially relevant articles.

The detailed search strategies are provided in supplementary **Appendix S1**. Results are reported according to the Preferred Reporting Items for Systematic Reviews and Meta-analyses (PRISMA) guidelines.^15^ This study is registered with PROSPERO.

### Data extraction

A full-text review was performed on the included articles and the following data were collected: total number of controls (when available), number of controls affected with the IBD risk factor, the total number of IBD patients (UC, CD, or both), and number of IBD patients affected with the risk factor. In addition to these data points, we also collected the study author, date, design, and country.

### Risk of Bias and Quality Assessment

Outliers were identified with externally standardized residuals in *R* version 3.6.1 (*R* Foundation for Statistical Computing). Absolute values of Z-scores greater than or equal to two were identified as outliers. Influential studies were identified as those with a Cook’s distance larger than three times the mean value. Studies that were outliers and influential were assessed for quality with the Newcastle-Ottawa Scale (NOS). The tool includes three domains: selection, comparability, and outcome/exposure. The threshold for good quality was a minimum of 3 stars in the selection domain and 1 star in the comparability domain and 2 stars in the outcome/exposure domain. Influential outliers that were good quality according to the NOS were included in the study.

### Model Development

The stepwise approach to the development of the computational model is shown in the supplementary **Table S1** and the final formula is given in ***Equation 1***. A Bayesian approach was chosen over a frequentist approach due to its ability to use informed priors to generate a probability that accounts for known background information. We used a Bayesian framework to calculate the probability of development or flare-up of IBD given the presence of IBD risk factors which were considered as multiple priors in Bayes’ formula. In the resulting *Equation 1* the risk factor prevalence was used as an equivalent of probability *(P)*: *IBD*, Inflammatory Bowel Disease (either Crohn’s disease or ulcerative colitis); *Pso*, Psoriasis; *HS*, Hidradenitis Suppurativa; *Smo*, Smoking status (current smoking for Crohn’s disease and former smoking for ulcerative colitis); *FDR*, first degree relative with IBD; *IL17i*, Interleukin-17 inhibitor. In Bayesian convention, the symbol “–” indicates the absence of a given factor and the vertical bar between probabilities (*X* | *Y*) symbolizes the probability of *X* given *Y* is true.

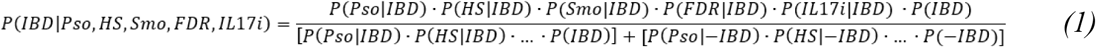

Two inputs were not available in the literature and thus required an expansion using Bayes’ theorem: *P(IL-17i*|*IBD)* and *P(FDR*|–*IBD)*. The proportion of individuals treated with IL-17i given the diagnosis of IBD, *P(IL-17i*|*IBD)*, can be calculated by inputting *P(IBD*|*IL17i), P(IBD)*, and *P(IL17i)* as shown in supplementary **Equation S1**. *P(IL17i)*, i.e. the probability of receiving IL-17i, is per definition 100% as all patients in our theoretical scenario will be receiving IL-17i. The other two inputs, *P(IBD)* and *P(IBD*|*IL17i)*, the prevalence of IBD among patients on an IL-17i, are reported in the literature. The second expanded input is *P(FDR*|–*IBD)*, as the prevalence of having a first-degree relative with IBD among people without a current diagnosis of IBD is not reported. The expansion of *P(FDR*|–*IBD)* is shown in supplementary **Equation S2**. *P(FDR)*, the probability of having a first degree relative with IBD in the general population, is a required input in the expansion of *P(FDR*|–*IBD)*. As *P(FDR)* is not reported in the literature, it was estimated using the prevalence of IBD,^7^ Canadian population of 37,058,856,^16^ and the Canadian fertility rate of 1.66^17^ as shown in supplementary **Equation S3**. This estimation was made assuming the child is the proband and that each family has a maximum of one affected FDR.

We considered the following scenarios when estimating the impact of IL17i on IBD risk: i) the baseline scenario compared psoriasis patient’s probability of IBD development (**Table S1**, Step 1a) with that of a psoriasis patient on IL-17i (**Table S1**, Step 1b), ii) the probability of IBD in a psoriasis patient with HS (**Table S1**, Step 2a) to a psoriasis patient with HS and on IL-17i (**Table S1**, Step 2b), iii) the probability of IBD in a psoriasis patient with a positive FDR (**Table S1**, Step 3a) to a psoriasis patient with a positive FDR and on IL-17i (**Table S1**, Step 3b), and iv) the probability of IBD in a psoriasis patient with a positive smoking status (**Table S1**, Step 4a) to a psoriasis patient with a positive smoking status on IL-17i (**Table S1**, Step 4b).

### Statistical analysis

Metaanalyses of the prevalence of risk factors were performed using *R* version 3.6.1. Proportions were first transformed using the double-arcsine method. These transformed proportions then underwent an inverse-variance weighted random-effects metaanalysis using the DerSimonian and Laird method.^18^ The double-arcsine transformation was chosen over logit transformation as it is better able to stabilize variances for proportions closer to 0 or 1. Logit transformations over-estimate variances for proportions close to 0 or 1 resulting in a small study with a prevalence around 0.5 outweighing a larger study with a smaller prevalence.^19^ We obtained pooled prevalences with 95% confidence intervals (CIs) of each IBD risk factor in CD and UC separately (**Table S7**). Heterogeneity was assessed using I^2^, which describes the total variance between studies due to heterogeneity of included studies^.20^

## Results

### Study selection

The literature search returned 29 studies that identified risk factors for IBD (supplementary **Fig. S2**). The following risk factors were identified: hidradenitis suppurativa (HS), psoriasis (Pso), first-degree relative with IBD (FDR), and smoking status (Smo, current smoking in CD and former smoking in UC).

### IBD and Psoriasis

We found 814 potentially relevant studies of which 7 studies^21–27^ were included in the meta-analysis (supplementary **Fig. S3**): one case-control, two cohort, and four cross-sectional studies. These studies included 28,883 total CD patients and 62,580 total UC patients (**Table S2**). A metaanalysis returned a pooled prevalence of psoriasis of 0.021 (95% CI: 0.015 – 0.029) and 0.014 (95% CI: 0.011 - 0.018) in CD and UC, respectively (**Table S7**). Using a general prevalence of CD of 0.096%^7^ and psoriasis of 2.0%^1^ combined with the pooled prevalence of psoriasis in CD (**Table S1**, Step 1a), the probability of CD or UC development given the presence of psoriasis (*P(CD*|*Psoriasis)* and *P(UC*|*Psoriasis)*) was in both instances 0.0010.

### IL-17i increase the risk of IBD in psoriasis patients

The MEDLINE search returned 369 studies from which 9 reports^28–34^ were included in the metaanalysis (supplementary **Fig. S4**). These nine studies referred to results from 29 clinical trials comprising 8,800 patients on an IL-17i (2,809 on ixekizumab, and 5,991 on secukinumab) for psoriasis, psoriatic arthritis, or ankylosing spondylitis (supplementary **Table S3**). Adverse events for these clinical trials were obtained from the US National Library of Medicine Clinical Trials Results Database. For two trials, FUTURE 2 and MEASURE 2, study data was not publicly available on the database and results were instead obtained from McInnes et al. 2015^35^ and Baeten et al. 2015^36^, respectively. The metaanalysis returned a pooled prevalence of IBD onset or flare in patients on IL-17i of 0.0019 (95% CI: 0.0013 – 0.0027) and 0.0022 (95% CI: 0.0015 - 0.0029) for CD and UC respectively (**Table S7**). Building on the previously calculated probability of CD and UC in psoriasis, the addition of IL-17 inhibition as a risk factor (**Table S1**, Step 1b) increased the probability of CD development from 0.0010 to 0.0037 and increased the probability of UC from 0.0010 to 0.0028 (**Table 1** and **Table 2**).

**Table 1.** Probability of Crohn’s disease (CD) in psoriasis patients stratified by IBD risk factors.^1^

| Risk Factor<br>(RF) | Control<sup>2</sup> |  |  | Patient on IL-17i<sup>2</sup> |  |  |
| --- | --- | --- | --- | --- | --- | --- |
|  | *P (CD \| Psoriasis, RF)*<br>(%) | Lower bound<br>(%) | Upper bound<br>(%) | *P (CD \| Psoriasis, IL-17i, RF)*<br>(%) | Lower bound<br>(%) | Upper bound<br>(%) |
| **No RF** | 0.10 | 0.07 | 0.14 | 0.37 | 0.18 | 0.69 |
| **FDR** | 36.14 | 18.99 | 53.39 | 53.56 | 24.89 | 76.08 |
| **Current**<br>**smoking** | 0.27 | 0.16 | 0.43 | 0.55 | 0.22 | 1.18 |
| **HS** | 5.93 | 2.69 | 10.98 | 11.38 | 3.76 | 25.52 |

<sup>1</sup>Abbreviations: CD, Crohn's disease; FDR, first degree relative with inflammatory bowel disease; HS, hidradenitis suppurativa; IL-17i, interleukin-17 inhibitor; RF, risk factor
<sup>2</sup>Upper and lower bounds calculated with lower and upper estimates of the prevalence of risk factors, respectively.

**Table 2.**
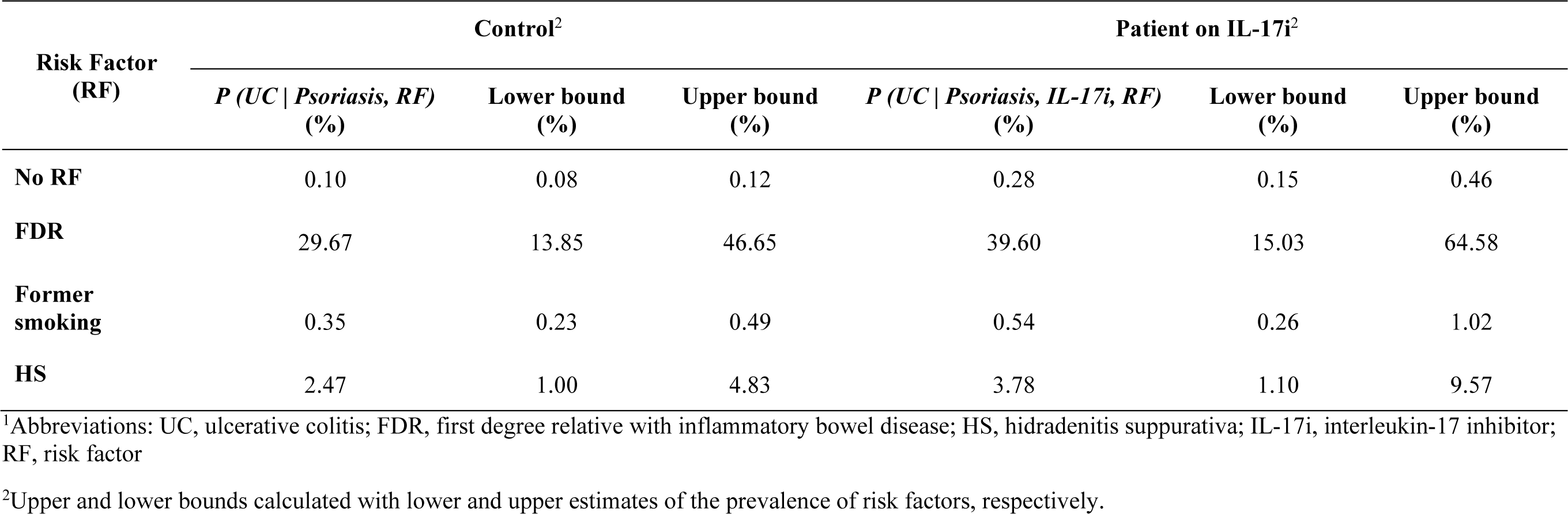
Probability of ulcerative colitis (UC) in psoriasis patients stratified by IBD risk factors.^1^

### Impact of risk factors on IBD in patients exposed to IL-17i

The literature search identified 3 risk factors associated with IBD: concomitant hidradenitis suppurativa (HS)^37–41^ (supplementary **Fig. S5** and **Table S4**), presence of the first-degree relative with IBD^42–57^ (supplementary **Fig. S6** and **Table S5**) and smoking^58–88^ (supplementary **Fig. S7, Table S6**). Metaanalyses produced values of pooled prevalences for each risk factor in CD and UC that were included in the Bayesian model (supplementary **Table S7**).

Bayesian modelling was done for both CD and UC to calculate the average, lower, and upper estimate of the probability of IBD development given risk factors using the average, lower, and upper bound estimates from the 95% confidence intervals for each risk factor, respectively (**Table 1** and **Table 2**). Finally, **Table 3** gives the values of the absolute risk increase of IBD attributed to the use of IL-17i. They allowed for calculating the number needed to harm (NNH) that reflects the number of psoriasis patients with a given risk factor profile who, if treated with IL-17i, would generate one case of CD or UC attributed to the use of the biologic.

**Table 3.** Increase in the absolute risk of Crohn’s disease (CD) or ulcerative colitis (UC) in a patient with different IBD risk factors exposed to IL-17i^1^

| Risk factor<br>*P*(IBD \| *RF*) | CD |  | UC |  |
| --- | --- | --- | --- | --- |
|  | AR Increase | NNH | AR Increase | NNH |
| *P*(IBD \| *Psoriasis*) | 0.27% | 373 | 0.18% | 564 |
| *P*(IBD \| *Psoriasis, Smoking*<sup>2</sup>) | 0.28% | 357 | 0.19% | 523 |
| *P*(IBD \| *Psoriasis, HS*) | 5.46% | 18 | 1.31% | 76 |
| *P*(IBD \| *Psoriasis, FDR*) | 17.43% | 6 | 9.92% | 10 |

<sup>1</sup>Abbreviations: AR, absolute risk; CD, Crohn’s disease; FDR, first-degree relative with IBD; HS, hidradenitis suppurativa, IBD, inflammatory bowel disease; NNH, number needed to harm attributed to IL-17i; UC, ulcerative colitis.
<sup>2</sup>Smoking: current smoking for CD and former smoking for UC.

### Study heterogeneity

The metaanalyses of proportions for both CD and UC in IL-17i showed low heterogeneities (I^2^=0%) and statistically insignificant p-values of (p<0.800) and (p<0.915) respectively. The remaining metaanalyses of psoriasis, smoking status, FDR, and HS in IBD showed high heterogeneities with I^2^ values greater than 75% and statistically significant p-values <0.001 (supplementary **Table S7**).

## Discussion

To our knowledge, this is the first study using Bayesian inference to quantify the probability of IBD flare or onset with IL-17i therapy. We confirmed that psoriasis is associated with both CD and UC. The association with CD seems to be stronger^89^ and our data showed that the prevalence of psoriasis in CD was 1.5 times greater than the prevalence of psoriasis in UC. Hitherto, the IBD risk in psoriasis has been done in terms of the relative change (relative risks, odds ratios or exposure-adjusted incidence rates) which is not intuitively understandable to the clinician. This created much debate on whether psoriasis patients should be screened for IBD before initiation of IL-17i therapy and what are the correct screening procedures. We have therefore focused on absolute risk which is recognized as a relevant, valid, and clinically useful measure included in the POEMs outcomes (Patient-Oriented Evidence that Matters).^90,91^ Especially, the use of the number needed to treat and the number needed to harm (NNH) as POEMs outcomes have been advocated and therefore adopted in our current study.

Our data clearly show that screening of asymptomatic psoriasis patients for IBD before initiation of IL-17i is not an efficient approach if the patient does not have any IBD risk factors. In those patients, the NNH necessary to see one case of IBD attributable to IL-17i is 373 (for CD) and 556 (for UC). The number needed to treat (NNT) to achieve PASI75 (1.18 for ixekizumab and 1.29 for secukinumab^92^) allow calculating the values of likelihood to be helped or harmed (LHH = NNH/NNT) to provide further perspective to the clinical value of IL17i. LLH values for IL17i indicate that the patient is ≅300 times more likely to benefit from the treatment than to be harmed.

However, clinical decision making should be modified for patients with preexisting IBD risk factors. Our metaanalysis uncovered three potential risk factors: smoking, family history (FDR), and co-morbid HS, of which only the two latter risk factors significantly affected NNH values. HS was very strongly associated with CD (58-fold increase in the probability of CD compared to a 25-fold increase of UC in psoriasis patients) and use of IL-17i in psoriasis patients with HS vastly increased the risk of IBD (NNH of 18 and 76 for CD and UC, respectively). Patients with HS have thus a ≅20-fold reduced LHH as compared with patients without IBD risk factors which could be used as an argument to avoid IL17i in those patients as the first-line antipsoriatic agent.

There is strong evidence suggesting a genetic basis for IBD. The polygenic nature of CD and UC was established with a genome search identifying susceptibility loci on chromosomes 12, 7, and 3. ^93^ First degree relatives of IBD patients have a 10-15-fold higher risk of developing IBD.^57,93^ Positive family history has been reported as the strongest recognizable risk factor for IBD,^94^ and was found to be the strongest risk factor in our study resulting in IL17i-related NNH of only 6 (for CD) and 10 (for UC).

This study is subject to limitations. Most importantly, we assumed independence between risk factors, which may not necessarily be the case for all risk factors, e.g. genetic predisposition to IBD and HS. Second, a significant proportion of the studies included were cross-sectional studies largely relying on the participants’ self-reporting of risk factors which may lead to recall bias. Third, there may be other risk factors yet to be discovered that would further improve our model.

In summary, the results of our study propose a simple approach to identify patients at increased risk for IBD in the context of IL-17i treatment. Asymptomatic patients who do not have any family history of IBD and do not have a comorbid HS can safely be treated with IL-17i as their added risk of inflammatory bowel disease is negligible and by far outweighed by the therapeutic benefit. In contrast, those who have one or both of those mentioned risk factors should be treated with caution and monitored for a new-onset IBD.

## Data Availability

Data used in this study is available in the supplementary materials.

